# Residual insecticide surface treatment for preventing malaria: a systematic review protocol

**DOI:** 10.1101/2021.12.13.21267747

**Authors:** Zachary Munn, Jennifer C. Stone, Timothy Hugh Barker, Carrie Price, Danielle Pollock, Alinune Nathanael Kabaghe, John E. Gimnig, Jennifer C. Stevenson

## Abstract

**Introduction:** Malaria presents a significant global public health burden, although substantial progress has been made, with vector control initiatives such as indoor residual surface spraying with insecticides and insecticide treated nets. There now exists many different approaches to apply residual insecticide to indoor and outdoor surfaces in malaria endemic settings. This review aims to synthesise the best available evidence regarding full or partial indoor or outdoor residual insecticide surface treatment for preventing malaria.

**Methods and Analysis:** This review will comprehensively search the literature (both published and unpublished) for any studies investigating the effectiveness of residual insecticide surface treatment for malaria. Studies will be screened to meet the inclusion criteria by a minimum of two authors, followed by assessment of risk of bias (using appropriate risk of bias tools for randomised and non-randomised studies) and extraction of relevant information using structured forms by two independent authors. Meta-analysis will be carried out where possible for epidemiological outcomes such as malaria, anaemia, malaria related mortality, all-cause mortality and adverse effects. Certainty in the evidence will be established with GRADE assessments.

**Ethics and Dissemination:** A full review report will be submitted to the Vector Control & Insecticide Resistance Unit, Global Malaria Program, WHO. A version of this report will be submitted for publication in an open access peer-reviewed journal. The report will inform the development of WHO recommendations regarding residual insecticide treatment for malaria. This systematic review does not require ethics approval as it is a review of primary studies.

**Registration:** PROSPERO, ID 293194 (in progress as of 24^th^ November, 2021)

## Background

### Description of the condition

Malaria is a parasitic, mosquito-borne, infectious disease that is highly transmissible, with a significant amount of the world’s population at risk.^1 2^ In 2020, there were an estimated 241 million cases of malaria worldwide with 627,000 deaths attributed to the disease.^2^ the World Health Organization (WHO) designated African region bears the largest burden in terms of malaria morbidity and mortality, with 228 million cases (approximately 95% of the global total) reported in 2020.^2^ Children under 5 years of age account for 77% of the total deaths.^2^

While malaria remains a significant global public health burden, substantial progress has been made since 2000 when global malaria deaths were estimated at 736,000. Global malaria incidence is estimated to have fallen from 81 cases per 1000 population at risk in 2000 to 59 per 1000 in 2020.^2^ Much of the decline in malaria burden was due to the scale up of malaria prevention and control interventions, particularly vector control such as insecticide treated nets (ITNs) and indoor residual spraying (IRS), which are estimated to have contributed to 68% and 11%, respectively, of the reduction in malaria burden between 2000 and 2015.^3^

### Description of the intervention

One of the primary measures of prevention and control of malaria includes residual insecticide surface treatment.^4^ This broad intervention may include the spraying (often described as indoor or outdoor residual spraying), painting or application of treated surface coverings (such as wallpaper) to the surface of walls and ceilings in houses, (potentially) both inside and outside of the home with insecticide to kill mosquitoes, reducing their longevity and population size. This may be full residual spraying (i.e. treating all surfaces) or partial (treating selected surfaces, rooms, parts of walls, etc). This practice, when applied indoors on internal walls and surfaces, has been used as an effective intervention in the control of malaria, by targeting *Anopheles* mosquitoes at rest, contributing to the significant reduction of malaria cases in multiple at-risk countries.^3 5-7^ A previous Cochrane review studying indoor residual spraying determined that while the primary evidence was limited in scope to determine efficacy, indoor residual spraying may be associated with overall health benefits.^8^

### Why it is important to do this review

While the global burden of malaria has declined since 2000, much of this was achieved between 2000 and 2015. Since 2015, malaria case incidence has declined by less than 2% with 31 countries reporting increased incidence of malaria.^1^ There is an urgent need to continue investigating potential measures for the control and prevention of malaria. A previous systematic review on this topic^8^ was limited to only randomized controlled trials (RCTs) and concluded that insufficient evidence existed at the time to determine efficacy. In addition, there have been no systematic reviews conducted on partial treatment of indoor surfaces or treatment of outdoor surfaces, or different application methods for insecticide (such as paints). Furthermore, there are now new insecticides and interventions that may have a different impact on malaria. Therefore, the current review aims to expand upon this earlier review, with broader inclusion of the various study designs, while also answering questions related to partial and outdoor surface treatment.

### Review Questions

1. In areas with ongoing malaria transmission, what are the relative benefits and harms of residual insecticide surface treatment where all surfaces of interior walls are treated compared with no residual insecticide surface treatment or other treatments/strategies to prevent malaria in adults and children?
2. In areas with ongoing malaria transmission, what are the relative benefits and harms of selective (or partial) residual insecticide surface treatment (such as where surfaces of interior walls are partially treated) compared with no residual insecticide surface treatment, full residual insecticide surface treatment, or other treatments/strategies to prevent malaria in adults and children?
3. In areas with ongoing malaria transmission, what are the relative benefits and harms of outdoor residual surface treatment (either stand alone or with concomitant treatment or indoor residual spraying) compared with no outdoor residual surface treatment or other treatments/strategies to prevent malaria in adults and children?

### Main objective

1. To assess the benefits (on malaria transmission or burden) and harms (adverse effects and unintended consequences) of residual insecticide surface treatments (either full or selective/partial indoor and outdoor residual surface treatment).

### Secondary objectives

1. To retrieve studies on contextual factors relating to residual insecticide surface treatments for preventing malaria. These factors include operational and feasibility considerations, such as area of intervention, coverage needed to optimize impact; acceptability; cost and resource use, cost effectiveness; impact on equity; values and preferences.
2. To retrieve entomological studies such as experimental hut studies and modelling as supportive data for interventions for which epidemiological outcomes may or may not have been reported.

## Methods

This review will be conducted in line with guidance from Cochrane ^9^, JBI ^10^ and GRADE.^11^ It will be reported in line with PRISMA 2020^12^ and this protocol is reported in line with PRISMA-P.^13^ The methods for the three review questions are presented together below. Where the methods differ based on the question they are presented separately. This review has been registered within PROSPERO, ID: 29314 (awaiting registration).

### Eligibility Criteria

#### Participants

Studies of adults and children who have resided in malaria endemic regions for more than one month (excluding travellers) are eligible for this review. Where studies have included travellers, attempts will be made to extract only information on residents, such as by contacting the authors or attempts to identify subgroups in the report. Where these attempts are unsuccessful, studies that combined data (residents and travellers) will be included only if more than 85% of participants were residents for more than one month.

#### Interventions

The interventions of interest are residual insecticide surface treatments, with surfaces being sprayed, painted or treated with insecticide, or application of insecticide-treated surface coverings such as wallpaper. There are no limitations on the type of insecticide that is studied. Specifically, the intervention of interest for the three review questions are:

1. Indoor residual insecticide surface treatment of the entirety of all indoor walls, and ceilings where applicable.
2. Selective/partial residual insecticide surface treatment (where insecticides are applied to only a limited surface area or limited wall application within the dwelling).
3. Outdoor residual insecticide surface treatment (either of the house/dwelling or other exterior surfaces of structures within a homestead)

For partial (or selective) treatment, the author’s definition of ‘partial’ will be used to categorise studies as opposed to a threshold established by the author team. In particular, we are interested in selective treatment that refers to applying an insecticide to parts of a wall, such as intentionally only spraying the bottom half of a wall. Different levels of houseful coverage will then be investigated through subgroup analysis. Where authors do not specify the household coverage, this will be presumed to be full surface treatment unless there is information to the contrary.

Non-residual, non-surface treatments (such as insecticide space sprays) will be excluded. The target of the interventions are mosquitoes at rest, and interventions aiming to limit mosquito movement or entry into a dwelling (such as curtains or nets) are not for consideration, but interventions such as wall paper and wall linings are relevant.

##### Background interventions

Background interventions (co-interventions) will likely be encountered and information on these will be extracted. These are interventions other than the intervention/s under consideration or any other malaria or vector-specific control intervention. Studies conducted where background interventions are present will be included as long as interventions (both malaria and non-malaria) were balanced between intervention and control arms.

#### Comparators

Comparators for the three review questions include no residual insecticide surface treatment as a comparator to full residual insecticide surface treatment, no residual insecticide surface treatment or full residual insecticide surface treatment as a comparator to selective or partial residual insecticide surface treatment, and no outdoor residual surface treatment as a comparator to outdoor residual surface treatment. All other comparators will be considered for inclusion, such as ITNs.

Reactive residual insecticide surface treatment, where treatment occurs in response in to an index case, will only be included in cases where it is a comparator to one of the three main interventions or where it is a background intervention.

#### Outcomes

The following outcomes were considered for inclusion and are grouped into epidemiological outcomes, entomological outcomes, unintended benefits and harms/ unintended consequences.

##### Epidemiological

- Malaria case incidence (symptomatic infection) in children and adults: defined as malaria symptoms with parasitological confirmation, such as with rapid diagnostic tests, thick/thin blood film for all species, PCR or ELISA, or other methods.
- Malaria infection incidence: active and passive parasitological confirmation with rapid diagnostic tests, thick/thin blood film, PCR or ELISA (both symptomatic and asymptomatic infection)
- Severe clinical malaria incidence: cerebral malaria or malaria with severe anaemia or based on site-specific definition with parasitological confirmation of malaria infection
- Malaria parasitaemia prevalence, confirmed using rapid diagnostic tests or thick/thin blood film for all species or PCR or other appropriate methods (both symptomatic and asymptomatic infection)
- Mild, moderate or severe anaemia prevalence defined by age- and sex-specific WHO haemoglobin cut-offs^14^
- Malaria mortality
- All-cause mortality

##### Entomological (Only when epidemiological outcomes have also been reported)

- Entomological inoculation rate (EIR), defined as the number of infective bites received by an individual during a season or annually
- Sporozoite rate, defined as the proportion of mosquitoes positive for sporozoites.
- Density of adult female *Anopeheles*, including human biting rate (number of mosquitoes caught per person or house per time period) and other measures of adult vector density.
- Mortality of adult female *Anopheles* measured by observation if it is knocked down, immobile or unable to stand or take off for 24 hours after exposure (or as reported in the primary evidence).
- Repellency of vector populations (e.g., anophelines) measured by contact irritancy and toxicity in the laboratory or through field evaluations.
- Parity rate, calculated as the number of parous females multiplied by 100 and divided by the total number of females dissected.

##### Unintended benefits

- Epidemiological impact on other vector-borne diseases

##### Harms and/or unintended consequences of interventions

- Adverse effects known to be associated with insecticides, including skin irritation, irritation of upper airways, nausea, and headache.
- Human behaviour changes e.g. proportion of time spent outside the house.
- Any influence on neighbouring houses e.g., increased vector abundance in unsprayed houses
- Environmental impacts such as biodiversity and ecosystem changes.
- Entomological impacts e.g., mosquito behaviour changes such as outdoor biting, biting times, feeding preference, development of insecticide resistance.

#### Setting

Studies conducted in settings with ongoing human malaria transmission. The presence of other background interventions will not impact on study eligibility. Studies where additional malaria interventions are considered standard of care were implemented will be included as long as interventions (both malaria and non-malaria) were balanced between intervention and control arms.

#### Study design

Randomized studies including cluster randomised controlled trials (no minimum amount of clusters), crossover trials and stepped wedge designs will be included.

Non-randomised study designs will be included when there is a comparison/control group present. This can include historical controls. There will be no exclusions on buffer period or length of intervention or timing of measurement of outcomes and these details will be extracted.

There will be no exclusions based on language or publication status (i.e. published, unpublished, in press, in progress, pre-print). There are no date limitations. For studies published in languages other than English, Google Translate will be used to determine whether the study meets inclusion criteria. Where studies are published in a language other than English and meet inclusion criteria, Google Translate translations will be reviewed by a person fluent in the language.

Due to the large amount of studies expected to be identified from the search, each article retrieved will be categorised based on their study methodology and design characteristics. Depending on the amount of studies identified for each intervention of interest, exclusions based on study design will be considered by the author team and the guideline development group and potentially implemented post searching, screening and retrieval of full text prior to extraction. This is to ensure the best available evidence to answer the research questions is used by optimising the signal to noise ratio from the evidence. This will allow the research team to prioritise and balance higher certainty evidence where it exists for a question against very low certainty evidence. For example, for the first intervention of interest (indoor residual insecticide surface treatment) where we expect to encounter the most studies, the group may choose to exclude from the level of before-and-after studies (with no control group) as it is unlikely the lower levels will provide higher certainty evidence. It will also be recorded whether studies have only one cluster/site/study area included in each arm or whether there are 2 or more within each arm. Some studies may contain elements of multiple study designs and as such the below are not mutually exclusive. Studies will be classified using the listing below:

1. Randomised controlled trials
2. Controlled trials
3. Controlled before and after studies
4. Interrupted time series (further split into studies where there are 3 or more data points pre/post the intervention and those where this is not the case)
5. Prospective/retrospective cohort studies (with exposed/non-exposed groups and baseline/post exposure data)
6. Before and after studies with no control group
7. Case control studies
8. Analytical cross-sectional studies at one time point
9. Descriptive cross-sectional studies at one time point
10. Modelling studies

### Search strategy

The search strategy aims to locate both published and unpublished studies and was developed with the input of a health librarian. An initial limited search of MEDLINE was undertaken to identify relevant articles on this topic. The terminology contained in the titles and abstracts of relevant articles, and the related subject headings and index terms used to describe the articles were used to develop a full search strategy for Ovid MEDLINE. This was also informed by a previous review on reactive indoor residual spraying and by using the searchrefinery tool.^15^ The search strategy, including all identified keywords and index terms, will be adapted for each included database and/or information source, by using Polyglot and with the aid of a health librarian.^16^ The reference list and citations of all studies undergoing extraction will be screened for additional studies using citationchaser ^17^ and/ or a related citations search.^18^ The full search strategies for major databases are available in <u>Supplementary file 1</u>.

The databases to be searched include Cochrane Central Register of Controlled Trials (CENTRAL), published in The Cochrane Library and including the Cochrane Infectious Diseases Group Specialized Register; MEDLINE (Ovid); EMBASE (Elsevier); CINAHL with full text (EBSCO), US National Institute of Health Ongoing Trials Register (www.ClinicalTrials.gov/); ISRCTN registry (www.isrctn.com/); The WHO’s International Clinical Trials Registry Platform (WHO ICTRP) (www.who.int.ictrp), the WHO Global Index Medicus (https://www.globalindexmedicus.net/), MSF publications and reports (www.msf.org), and Demographics and Health Surveys (DHS) indicator surveys and reports (dhsprogram.com).

Additionally, experts in the field and relevant organisations will be asked whether they know of any studies (completed or ongoing) that are relevant to this review topic. Organisations included the Malaria Elimination Initiative at the University of California at San Francisco the Malaria Eradication Research Agenda Consortium, the Malaria Eradication Scientific Alliance, and the Bill and Melinda Gates Foundation.

Relevant conference proceedings will also be reviewed (where accessible) for potential studies. This will include searching the most recent conferences for the Multilateral Initiative on Malaria Pan-African Malaria Conference, American Society of Tropical Medicine and Hygiene, and the Malaria in Melbourne event.

### Study selection and screening

Following the search, all identified citations will be collated and uploaded into EndNote™ and duplicates removed. The studies will then be imported into Covidence for screening, with titles and abstracts screened by two or more independent reviewers for assessment against the inclusion criteria for the review. Potentially relevant studies will be retrieved in full. The full text of selected citations will be assessed in detail against the inclusion criteria by two or more independent reviewers in Covidence. Reasons for exclusion of studies at full text that do not meet the inclusion criteria will be recorded and reported. Any disagreements that arise between the reviewers at each stage of the selection process will be resolved through discussion, or with an additional reviewer/s. The results of the search and the study inclusion process will be reported in full in the final review and presented in a Preferred Reporting Items for Systematic Reviews and Meta-analyses (PRISMA 2020) flow diagram.^12^ Where relevant systematic reviews are identified in the search, we will review the list of included and excluded studies for consideration, however the systematic reviews themselves will not be included in our review.

Where members of the author team are named as authors on primary studies identified through the search, they will be excluded from any decision-making regarding the inclusion of the study or the assessment of the study risk of bias.

#### Contextual Factors and Entomological Studies

During the screening process, studies that potentially provide important information regarding the contextual factors required to make a recommendation by a guideline panel will be tagged within Covidence at the full text screening stage. These studies, if they do not provide data relating to the outcomes of interest in this review, will be retrieved and collated for further consideration in the future (i.e. data extraction and synthesis), potentially as subsequent standalone reviews. There are no restrictions based on the study design type for contextual factors. Likewise, entomological studies will be retrieved and collated for further consideration.

### Data extraction

Data will be extracted from papers included in the review by two or more independent reviewers using a tailored data extraction tool developed by the reviewers. The data extracted will include specific details about the participants, concept, context, study methods and key findings relevant to the review question/s. The full list of items to be extracted is included in <u>Supplementary file 2</u>. Any disagreements that arise between the reviewers will be resolved through discussion, or with an additional reviewer/s. If appropriate, authors of papers will be contacted to request missing or additional data where required. The data extraction form for study characteristics and outcome data will be piloted on two studies prior to finalisation.

### Assessment of the Risk of Bias

Two review authors will independently assess the risk of bias for each study using the Cochrane Risk of Bias 2 tool for randomised controlled trials (and the Risk of Bias 2 tool for cluster trials where appropriate)^19^ and the ROBINS-I for non-randomised studies,^20^ using the templates available at riskofbias.info. Any disagreements that arise between the reviewers will be resolved through discussion, or with an additional reviewer/s. Authors of papers will be contacted to request missing or additional data, where required. Risk of bias will be undertaken at the result level.

### Data synthesis and Meta-analysis

#### Meta-analysis

We will pool studies, where possible, in a meta-analysis using Review Manager 5 (RevMan5)^21^ where two or more studies report results for the same outcome in a format conducive to meta-analysis. Where there is only one study contributing data to a particular outcome for a comparison, a forest plot will still be presented for illustrative purposes (without a meta-analysis estimate). A narrative synthesis of the results will accompany any meta-analysis or present the results in the absence of a meta-analysis. Results from randomised controlled trials and non-randomised controlled trials will be analysed separately.

For dichotomous data we will calculate effect sizes as relative risks and present these with 95% confidence intervals (CIs). When there are no events in a treatment arm, RevMan will add a fixed value of 0.5 to the empty cell. If there are no events in the study, the study will not contribute to the pooled relative estimate of effect from the meta-analysis, however we will keep these results to inform baseline risk for absolute as opposed to relative comparisons and use risk difference instead of relative risk. Where incidence rates are reported and appropriate, we will calculate the incidence rate ratios. Where possible, adjusted estimates will be extracted for synthesis from non-randomised studies. A random effects model will be used when there are more than two studies and a fixed effect model when there are only two studies.

#### Assessment of heterogeneity and publication biases

We will assess clinical and methodological heterogeneity by evaluating the similarities and differences across studies in terms of the population, the intervention and study design. We will assess statistical heterogeneity visually by inspection of the forest plot and statistically by Cochran’s Q (P value 0.05), and by I², which is a statistic used for quantifying inconsistency in meta-analysis. In a meta-analysis, which uses a random-effects model, we will also report tau^2^, an estimate of between-study variability.

We will interpret the I² according to the guidance in the Cochrane Handbook for Systematic Reviews of Interventions,^22^ bearing in mind the limitations of concrete thresholds for I²:

1. 0% to 40%: might not be important;
2. 30% to 60%: may represent moderate heterogeneity;
3. 50% to 90%: may represent substantial heterogeneity; or
4. 75% to 100%: considerable heterogeneity.

To address publication bias, we will seek both published and unpublished literature. Additionally, if there is an adequate number of studies (at least 10), we will create funnel plots to investigate reporting bias. ^23^ In the case of 10 or more studies, for continuous data, we will use Egger’s test. ^24^ If effect sizes appear to depend on the size of the trial, we will explore whether this association is due to heterogeneity or publication bias.

#### Unit of analysis issues

We are likely to encounter unit of analysis issues in this systematic review, as groups of individuals are likely to be randomised and assigned together in clusters.^25^ We will take appropriate measures to address unit of analysis issues such as using the generic inverse variance method in RevMan when studies have analysed their data accounting for their cluster design. It is likely that authors will have taken steps to control for clustering in their analysis, however, if not, we will inflate standard errors following the methods in the Cochrane Handbook.^25^

#### Subgroup and sensitivity analysis

A number of potential treatment effect modifiers have been identified which will be assessed using subgroup analysis where data is available. These will also be explored as potential contributors to heterogeneity in the overall analysis. These include:

- Coverage / deployment strategy of intervention applied e.g. number of rounds of spraying/treatment per year/season, timing of outcome measurement.
- Insecticide used and class (i.e. organochlorines such as DDT, pyrethroids such as deltamethrin, carbamates such as bendiocarb, organophosphates such as pirimiphos-methyl, neonicotinoids such as clothianidin, and pyrroles such as chlorfenapyr).
- Target sites, mode of action, and duration required to produce such effect(s).
- Dwelling material and type (e.g., fixed versus temporary/ humanitarian structures)
- Level of household coverage (i.e. different amounts of ‘partial’ treatment or label indications (bedrooms only vs all house; 1 metre from the floor vs full, 10cm from the ceiling vs full).
- Level of transmission; seasonality of transmission (High: incidence of about 450 cases/1000 persons/year or Plasmodium falciparum (Pf) / Plasmodium vivax (Pv) prevalence of >=35%; Moderate: incidence of 250-450 per 1000 persons per year and Pf/Pv prevalence of 10-35%; Low: incidence of 100-250 per 1000 persons per year and Pf/Pv prevalence of 1-10%; Very low: incidence of <100 per 1000 persons per year and Pf/Pv prevalence <1%.) (the level of transmission will be categorized according to the schema found in the *Framework for malaria elimination)*.^26^
- Coverage of other background interventions.
- Vector species characteristics e.g., species, behaviours, insecticide resistance, etc.
- Setting e.g., rural/ urban/ peri-urban.
- Population demographics e.g., sex/age/SES/ethnicity etc.
- Human behaviour e.g., sleeping behaviour, replastering of houses.
- Time from implementation.
- Subgroups with/without ITNs (in addition to further subgroups if possible for pyrethroid-only nets and pyrethroid-PBO nets).

Sensitivity analyses will be conducted to determine the following:

1. The impact of bias by excluding studies that are at a high risk of bias. If there is no difference between the high risk of bias and low risk of bias studies, the original analysis result will stand. In the case where there are differences between the estimate of the pooled high risk of bias studies as compared to the low risk of bias studies, all the results will be presented and the preference of the guideline group will be followed as to what estimate (i.e. the full analysis or only the low risk of bias studies) should be used for grading and as the basis of recommendations.
2. Where we have inflated standard errors for trials where cluster designs have not been taken into account, we will analyse trials as if the individual was the unit of randomisation.

### GRADE

The Grading of Recommendations, Assessment, Development and Evaluation (GRADE) approach ^11^ for grading the certainty of evidence will be followed and GRADE Evidence Profiles will be created using GRADEpro GDT for each comparison. Evidence from RCTs and non-RCTs will all start off as high certainty. Historically in the GRADE approach, evidence from non-randomized studies started as low certainty but as we are using ROBINS-I for assessing risk of bias^27^ they start off as high. The evidence profile will present the following information where appropriate: absolute risks for the treatment and control, estimates of relative risk, and a rating of the certainty of the evidence based on the risk of bias, indirectness, heterogeneity, imprecision and risk of publication bias of the review results. Assessments from RCTs and non-RCTS will be presented separately. The outcomes reported in the evidence profiles will be:

- Malaria case incidence (symptomatic infection) by species (i.e., *P. falciparum, P. malariae, P. vivax, P. ovale, and P. knowlesi*)
- Malaria infection incidence by species (i.e., *P. falciparum, P. malariae, P. vivax, P. ovale, and P. knowlesi*)
- Malaria parasitaemia prevalence (both symptomatic and asymptomatic infection) by species (i.e., *P. falciparum, P. malariae, P. vivax, P. ovale, and P. knowlesi*)
- Anaemia prevalence
- Malaria mortality
- All-cause mortality
- Adverse effects known to be associated with insecticides, including skin irritation, irritation of upper airways, nausea, and headache.

## Data Availability

All data from the study (including extraction details, risk of bias assessments, statistical files [such as RevMan files]) will be made available via a publically available project space using open science platforms. A full review report will be submitted to the Vector Control & Insecticide Resistance Unit, Global Malaria Program, WHO. A version of this report will be submitted for publication in an open access peer-reviewed journal. The report will inform the development of WHO recommendations regarding residual insecticide surface treatment for malaria.

## Ethics and Dissemination

This systematic review does not require ethics approval as it is a review of primary studies. We will not collect any identified individual level participant data within this study. All data from the study (including extraction details, risk of bias assessments, statistical files [such as RevMan files]) will be made available via a publically available project space using open science platforms. A full review report will be submitted to the Vector Control & Insecticide Resistance Unit, Global Malaria Program, WHO. A version of this report will be submitted for publication in an open access peer-reviewed journal. The report will inform the development of WHO recommendations regarding residual insecticide surface treatment for malaria.

## Consent for publication

Publication of this protocol was formally cleared through the World Health Organization’s approval system for external publications.

## Acknowledgements

The authors would like to acknowledge and thank staff from the Global Malaria Programme, WHO, for their input and support. We would like to acknowledge Jan Kolaczinski Head, Vector Control & Insecticide Resistance Unit, Global Malaria Program, WHO for Department of Internal Medicine, and Elie Akl, American University of Beirut, Lebanon for their guidance and feedback. We would finally like to acknowledge the feedback from the members of the Guideline Development Group for malaria vector control for this project.

## Author contributions

JCS and colleagues conceptualised the work.

ZM, JS, TB authored first draft of the protocol

CP led the development of the search strategy.

JCS collected and summarised stakeholder feedback on the protocol.

All authors reviewed the content, commented on the methods, provided feedback, contributed to drafts and approved the content.

## Funding

This work is funded by the World Health Organisation, APW202747274.

## Disclaimer

The findings and conclusions in this report are those of the author(s) and do not necessarily represent the official position of the Centers for Disease Control and Prevention or the Department of Health and Human Services nor that of the World Health Organization.

## Notes

Zachary Munn is employed by JBI, an evidence-based healthcare research and development organisation situated within the University of Adelaide and is supported by an NHMRC Investigator Grant 1195676.

### Competing Interest Statement

This work is funded by the World Health Organisation, APW202747274.
Zachary Munn is employed by JBI, an evidence-based healthcare research and development organisation situated within the University of Adelaide and is supported by an NHMRC Investigator Grant 1195676.
The findings and conclusions in this report are those of the author(s) and do not necessarily represent the official position of the Centers for Disease Control and Prevention or the Department of Health and Human Services nor that of the World Health Organization.

