## Supplementary file 1 for "Residual insecticide surface treatment for preventing malaria: a systematic review protocol"

| Databases | Cochrane Library, MEDLINE (Ovid), Embase (Elsevier), CINAHL with Full Text (EBSCO), Scopus, WHO Global Index Medicus |
| --- | --- |
| Registries | ClinicalTrials.gov, ISRCTN Registry, WHO ICTRP |
| Additional Sources | MSF Publications and Reports |
| Date Run |  |
| Total Results |  |
| Duplicates Removed |  |
| Unique Results |  |
| Searcher(s) | - Carrie Price, MLS, Health Professions Librarian, Towson University, Towson, Maryland, USA |

| Line | Ovid MEDLINE | Results |
| --- | --- | --- |
| 1 | exp malaria/ |  |
| 2 | (black water fever OR black water fevers OR blackwater fever OR blackwater fevers OR malaria* OR marsh fever OR marsh fevers OR paludism OR plasmodia OR plasmodiosis OR plasmodium OR plasmodiums OR remittent fever OR remittent fevers OR swamp fever OR swamp fevers).mp. |  |
| 3 | 1 OR 2 |  |
| 4 | (indoor OR in door OR outdoor OR out door OR indoors OR outdoors OR in doors OR out doors OR interior OR exterior OR interiors OR exteriors OR inside OR outside OR insides OR outsides).mp. |  |
| 5 | (chair OR chairs OR counter* OR door OR doorknob* OR doors OR dwelling OR dwellings OR furniture* OR home OR homes OR homestead OR homesteads OR house OR houses OR household OR households OR hut OR huts OR residence OR residences OR paint OR paints OR plaster* OR shelf OR shelves OR surface OR surfaces OR table OR tables OR tabletop OR tabletops OR wall OR walls OR residual*).mp. |  |
| 6 | (additive OR additives OR aerosol* OR fumigant* OR fumigat* OR IRS OR spray* OR treatment OR treatments).mp. |  |
| 7 | 4 AND 5 AND 6 |  |
| 8 | 3 AND 7 |  |

Date run:

Total Result:

| Line | Embase | Results |
| --- | --- | --- |
| 1 | 'malaria'/exp |  |
| 2 | ('black water fever' OR 'black water fevers' OR 'blackwater fever' OR 'blackwater fevers' OR malaria* OR 'marsh fever' OR 'marsh fevers' OR 'paludism' OR 'plasmodia' OR 'plasmodiosis' OR 'plasmodium' OR 'plasmodiums' OR 'remittent fever' OR 'remittent fevers' OR 'swamp fever' OR 'swamp fevers'):ti,ab,kw |  |
| 3 | 1 OR 2 |  |
| 4 | ('indoor' OR 'in door' OR 'outdoor OR 'out door' OR 'indoors' OR 'outdoors' OR 'in doors' OR 'out doors' OR 'interior' OR 'exterior' OR 'interiors' OR 'exteriors' OR 'inside' OR 'outside' OR 'insides' OR 'outsides'):ti,ab,kw |  |
| 5 | ('chair' OR 'chairs' OR counter* OR 'door' OR doorknob* OR 'doors' OR 'dwelling' OR 'dwellings' OR furniture* OR 'home' OR 'homes' OR 'homestead' OR 'homesteads' OR 'house' OR 'houses' OR 'household' OR 'households' OR 'hut' OR 'huts' OR 'residence' OR 'residences' OR 'paint' OR 'paints' OR 'plaster*' OR 'shelf' OR 'shelves' OR 'surface' OR 'surfaces' OR 'table' OR 'tables' OR 'tabletop' OR 'tabletops' OR 'wall' OR 'walls' OR residual*):ti,ab,kw |  |
| 6 | (additive' OR 'additives' OR aerosol* OR fumigant* OR fumigat* OR 'IRS' OR spray* OR 'treatment' OR 'treatments'):ti,ab,kw |  |
| 7 | 4 AND 5 AND 6 |  |
| 8 | 'indoor residual spraying'/exp |  |
| 9 | 7 OR 8 |  |
| 10 | 3 AND 9 |  |

| Line | Cochrane Library | Results |
| --- | --- | --- |
| 1 | [mh "malaria"] |  |
| 2 | ("black water fever" OR "black water fevers" OR "blackwater fever" OR "blackwater fevers" OR "malaria*" OR "marsh fever" OR "marsh fevers" OR "paludism" OR "plasmodia" OR "plasmodiosis" OR "plasmodium" OR "plasmodiums" OR "remittent fever" OR "remittent fevers" OR "swamp fever" OR "swamp fevers"):ti,ab,kw |  |
| 3 | #1 OR #2 |  |
| 4 | ("indoor" OR "in door" OR "outdoor" OR "out door" OR "indoors" OR "outdoors" OR "in doors" OR "out doors" OR "interior" OR "exterior" OR "interiors" OR "exteriors" OR "inside" OR "outside" OR "insides" OR "outsides"):ti,ab,kw |  |
| 5 | ("chair" OR "chairs" OR counter* OR "door" OR doorknob* OR "doors" OR "dwelling" OR "dwellings" OR furniture* OR "home" OR "homes" OR "homestead" OR "homesteads" OR "house" OR "houses" OR "household" OR "households" OR "hut" OR "huts" OR "residence" OR "residences" OR "paint" OR "paints" OR plaster* OR "shelf" OR "shelves" OR "surface" OR "surfaces" OR "table" OR "tables" OR "tabletop" OR "tabletops" OR "wall" OR "walls" OR residual*):ti,ab,kw |  |
| 6 | ("additive" OR "additives" OR aerosol* OR fumigant* OR fumigat* OR "IRS" OR spray* OR "treatment" OR "treatments"):ti,ab,kw |  |
| 7 | #4 AND #5 AND #6 |  |
| 8 | #3 AND #7 |  |

| Line | Scopus (use advanced document search) | Results |
| --- | --- | --- |
| 1 | TITLE-ABS-KEY({black water fever} OR {black water fevers} OR {blackwater fever} OR {blackwater fevers} OR malaria* OR {marsh fever} OR {marsh fevers}OR {paludism} OR {plasmodia} OR {plasmodiosis} OR {plasmodium} OR {plasmodiums} OR {remittent fever} OR {remittent fevers} OR {swamp fever} OR {swamp fevers}) |  |
| 2 | TITLE-ABS-KEY ({indoor} OR {in door} OR {outdoor} OR {out door} OR {indoors} OR {outdoors} OR {in doors} OR {out doors} OR {interior} OR {exterior} OR {interiors} OR {exteriors} OR {inside} OR {outside} OR {insides} OR {outsides}) |  |
| 3 | TITLE-ABS-KEY ({chair} OR {chairs} OR counter* OR {door} OR doorknob* OR {doors} OR {dwelling} OR {dwellings} OR furniture* OR {home} OR {homes} OR {homestead} OR {homesteads} OR {house} OR {houses} OR {household} OR {households} OR {hut} OR {huts} OR {residence} OR {residences} OR {paint} OR {paints} OR plaster* OR {shelf} OR {shelves} OR {surface} OR {surfaces} OR {table} OR {tables} OR {tabletop} OR {tabletops} OR {wall} OR {walls} OR residual*) |  |
| 4 | TITLE-ABS-KEY ({additive} OR {additives} OR aerosol* OR fumigant* OR fumigat* OR {IRS} OR spray* OR {treatment} OR {treatments}) |  |
| 5 | #2 AND #3 AND #4 |  |
| 6 | #1 AND #5 |  |

| Line | CINAHL with full text | Results |
| --- | --- | --- |
| 1 | MH "malaria" |  |
| 2 | ("black water fever" OR "black water fevers" OR "blackwater fever" OR "blackwater fevers" OR malaria* OR "marsh fever" OR "marsh fevers" OR "paludism" OR "plasmodia" OR "plasmodiosis" OR "plasmodium" OR "plasmodiums" OR "remittent fever" OR "remittent fevers" OR "swamp fever" OR "swamp fevers") |  |
| 3 | 1 OR 2 |  |
| 4 | ("indoor" OR "in door" OR "outdoor" OR "out door" OR "indoors" OR "outdoors" OR "in doors" OR "out doors" OR "interior" OR "exterior" OR "interiors" OR "exteriors" OR "inside" OR "outside" OR "insides" OR "outsides") |  |
| 5 | ("chair" OR "chairs" OR counter* OR "door" OR doorknob* OR "doors" OR "dwelling" OR "dwellings" OR furniture* OR "home" OR "homes" OR "homestead" OR "homesteads" OR "house" OR "houses" OR "household" OR "households" OR "hut" OR "huts" OR "residence" OR "residences" OR "paint" OR "paints" OR plaster* OR "shelf" OR "shelves" OR "surface" OR "surfaces" OR "table" OR "tables" OR "tabletop" OR "tabletops" OR "wall" OR "walls" OR residual*) |  |
| 6 | ("additive" OR "additives" OR aerosol* OR fumigant* OR fumigat* OR "IRS" OR spray* OR "treatment" OR "treatments") |  |
| 7 | 4 AND 5 AND 6 |  |
| 8 | 3 AND 7 |  |

| Line | WHO Global Index Medicus (use advanced search)  <https://pesquisa.bvsalud.org/gim/advanced/?lang=en> | Results |
| --- | --- | --- |
| 1 | (tw:(("black water fever" OR "black water fevers" OR "blackwater fever" OR "blackwater fevers" OR malaria* OR "marsh fever" OR "marsh fevers" OR "paludism" OR "plasmodia" OR "plasmodiosis" OR "plasmodium" OR "plasmodiums" OR "remittent fever" OR "remittent fevers" OR "swamp fever" OR "swamp fevers"))) AND (tw:( ("indoor" OR "in door" OR "outdoor" OR "out door" OR "indoors" OR "outdoors" OR "in doors" OR "out doors" OR "interior" OR "exterior" OR "interiors" OR "exteriors" OR "inside" OR "outside" OR "insides" OR "outsides") AND ("chair" OR "chairs" OR counter* OR "door" OR doorknob* OR "doors" OR "dwelling" OR "dwellings" OR furniture* OR "home" OR "homes" OR "homestead" OR "homesteads" OR "house" OR "houses" OR "household" OR "households" OR "hut" OR "huts" OR "residence" OR "residences" OR "paint" OR "paints" OR plaster* OR "shelf" OR "shelves" OR "surface" OR "surfaces" OR "table" OR "tables" OR "tabletop" OR "tabletops" OR "wall" OR "walls" OR residual*) AND ("additive" OR "additives" OR aerosol* OR fumigant* OR fumigat* OR "IRS" OR spray* OR "treatment" OR "treatments"))) |  |

| ClinicalTrials.gov Searches | Results |
| --- | --- |
| Condition or disease: Malaria  Other Terms: spraying |  |
| Condition or disease: Malaria  Other Terms: IRS |  |
| Condition or disease: Malaria  Other terms: residual |  |
| TOTAL |  |

| ISRCTN registry | Results |
| --- | --- |
| Malaria |  |

| WHO ICTRP  <https://trialsearch.who.int/Default.aspx> | Results |
| --- | --- |
| Malaria AND spraying |  |
| Malaria AND IRS |  |
| Malaria AND residual |  |
| TOTAL |  |

| MSF Publications and Reports | Results |
| --- | --- |
