## Supplementary file 2 for "Residual insecticide surface treatment for preventing malaria: a systematic review protocol"

Data Extraction Form

| **Study characteristics (Study reference)** |
| --- |
| **DESIGN** |
| **Lead Author** |
| **Publication year** |
| **Time period study was conducted** |
| **Design** |
| **Method of Randomization/ matching or other** |
| **Unit of allocation** |
| **Number of units** |
| **Adjustment for clustering** |
| **Outcomes assessed** |
| **SETTING** |
| **Country** |
| **Site/s (town/settlements/region)** |
| **Seasonality of transmission** |
| **Level of transmission** |
| **Study site (e.g., rural/ urban/ peri-urban/ level of urbanicity)** |
| **Vector species and vector profile details (i.e., behaviours, resistance profile, parity, sporozoite rates and all other reported information)** |
| **Malaria species** |
| **PARTICIPANTS (those who received the intervention and on whom impact was measured)** |
| **Total trial participants in each group/arm (including number of exclusions and reasons)** |
| **Number of clusters per group/arm** |
| **Participants who received the intervention (if provided) and participants on whom the impact was measured** |
| **Recruitment method** |
| **Recruitment rates** |
| **Eligibility criteria** |
| **Characteristics and Demographics**  **(age, sex, ethnicity, SES, time as resident, and all other reported demographics)** |
| **Frequency of travel in the last month** |
| **Cluster details including buffer sizes between clusters, other indication of dilution effects** |
| **INTERVENTION and COMPARISON** |
| **Insecticide brand, type, formulation (including active ingredient), dose, duration (this includes timing and frequency of application)** |
| **Insecticide application method(s) and strategy e.g., spray, paint, treated materials, wallpaper)** |
| **Personnel applying the intervention** |
| **Target area (inside/outside)** |
| **Coverage of household (full, selective/partial)** |
| **Coverage across cluster/site/jurisdiction** |
| **Length of intervention, including number of rounds of spraying/application per year/ season** |
| **Type of dwelling (e.g., fixed or temporary) and construction material of surfaces insecticides applied to (e.g., cement, brick or mud walls, canvas etc)** |
| **Time to start intervention after index case** |
| **Human behaviour (e.g., sleeping behaviour, re-plastering of houses)** |
| **Details of Comparison** |
| **How intervention and comparison was measured** |
| **Coverage across cluster/site/jurisdiction for the comparison** |
| **Background interventions**  **(all reported in primary study – e.g., spraying prior to treatment period and other malaria or vector-specific control interventions [indoor surface treatment, nets/ other insecticides/ barrier], cointerventions, treatment of individuals that may impact outcome)** |
| **Costs** |
| **Resources needed/ used** |
| **OUTCOME X (repeat as necessary for each outcome)** |
| **Name / Definition** |
| **Assessment Metric** |
| **Events** |
| **Total (or unit time)** |
| **Time of outcome assessment** |
| **Results**  **(i.e., main results – epidemiological and/ or entomological, unadjusted and adjusted, secondary outcomes, subgroups, and clusters)** |
| **Effect type** |
| **Unintended benefits** |
| **Harms** |
| **ADDITIONAL DATA** |
| **Other contextual information present/measured/reported) (feasibility, acceptability, preferences/values, impact on equity)** |
| **Entomological outcomes measured (list)** |
| **Other** |
| **Source of funding** |
| **Possible conflicts of interest** |
